# Prospective Evaluation of a Breast Cancer Risk Model Integrating Classical Risk Factors and Polygenic Risk in 15 Cohorts from Six Countries

**DOI:** 10.1101/19011171

**Authors:** Amber N Wilcox, Parichoy Pal Choudhury, Chi Gao, Anika Hüsing, Mikael Eriksson, Min Shi, Christopher Scott, Brian D Carter, Kara Martin, Elaine Harkness, Mark N Brook, Thomas U Ahearn, Nasim Mavaddat, Antonis C Antoniou, Jenny Chang-Claude, Jacques Simard, Michael E Jones, Nick Orr, Minouk J Schoemaker, Anthony J Swerdlow, Sarah Sampson, William G Newman, Elke M van Veen, D. Gareth R Evans, Robert J MacInnis, Graham G Giles, Melissa Southey, Roger L Milne, Susan M Gapstur, Mia M Gaudet, Stacey J Winham, Kathy Brandt, Aaron Norman, Celine M Vachon, Dale P Sandler, Clarice R Weinberg, Kamila Czene, Marike Gabrielson, Per Hall, Carla H van Gils, Kay-Tee Khaw, Myrto Barrdahl, Rudolf Kaaks, Paul M Ridker, Julie E Buring, Dan I Chasman, Douglas F Easton, Marjanka K Schmidt, Peter Kraft, Montserrat Garcia-Closas, Nilanjan Chatterjee

## Abstract

**PURPOSE:** Risk-stratified breast cancer prevention requires accurate identification of women at sufficiently different levels of risk. We conducted a comprehensive evaluation of a model integrating classical risk factors and a recently developed 313-variant polygenic risk score (PRS) to predict breast cancer risk.

**METHODS:** Fifteen prospective cohorts from six countries with 237,632 women (7,529 incident breast cancer patients) of European ancestry aged 19-75 years at baseline were included. Calibration of five-year risk was assessed by comparing predicted and observed proportions of cases overall and within risk categories. Risk stratification for women of European ancestry aged 50-70 years in those countries was evaluated by the proportion of women and future breast cancer cases crossing clinically-relevant risk thresholds.

**RESULTS:** The model integrating classical risk factors and PRS accurately predicted five-year risk. For women younger than 50 years, median (range) expected-to-observed ratio across the cohorts was 0.94 (0.72 to 1.01) overall and 0.9 (0.7 to 1.4) at the highest risk decile. For women 50 years or older, these ratios were 1.04 (0.73 to 1.31) and 1.2 (0.7 to 1.6), respectively. The proportion of women in the general population identified above the 3% five-year risk threshold (used for recommending risk-reducing medications in the US) ranged from 7.0% in Germany (∼841,000 of 12 million) to 17.7% in the US (∼5.3 of 30 million). At this threshold, 14.7% of US women were re-classified by the addition of PRS to classical risk factors, identifying 12.2% additional future breast cancer cases.

**CONCLUSION:** Evaluation across multiple prospective cohorts demonstrates that integrating a 313-SNP PRS into a risk model substantially improves its ability to stratify women of European ancestry for applying current breast cancer prevention guidelines.

## INTRODUCTION

Clinical guidelines for breast cancer prevention and early detection use risk thresholds to identify women eligible for interventions, e.g., genetic counseling, tailored screening, risk-reducing drugs or surgery.^1-3^ Risk assessment under these guidelines can be based on age, family history of cancer, mutation status for breast cancer genes such as *BRCA1* and *BRCA2*, history of benign breast disease, and other risk factors. Several risk models are available to estimate absolute risk, each including different sets of risk factors and aimed at different clinical scenarios.^4,5^ For instance, family-based models using pedigree-level information on family history of cancer and mutation testing are typically used for genetic counseling of women in cancer genetics clinics, whereas models with more limited family history information and personal risk factors are often used to identify women for referral to genetic or oncology clinics. Many of these models are used clinically with limited evidence on their comparative performance across populations.^6-8^ Additionally, there is a need to improve the risk discrimination of current models, particularly those assessing risk in the general population.

Polygenic risk scores (PRS) based on common single nucleotide polymorphisms (SNPs), identified by genome-wide association studies (GWAS)^9^ can provide improved risk stratification of breast cancer, i.e., allow accurate identification of higher proportion of populations at the extremes, compared to other risk factors for women with or without family history.^8,10-14^ Risk assessment using PRS is already commercially available and marketed to clinicians.^15,16^ However, while PRSs alone have been demonstrated to provide accurate relative risks for breast cancer in women of European ancestry,^17^ evidence on the accuracy of predicted risk from models integrating PRS and classical risk factors in prospective cohort studies is very limited.^7,8,11,16,18^ Thus, there is an urgent need for this type of assessment in large cohorts across multiple populations.

Individualized Coherent Absolute Risk Estimator (iCARE) is a new flexible tool^19^ to develop and validate risk prediction models in the general population. The iCARE models for breast cancer based on classical risk factors showed similar or better prediction accuracy and discrimination compared to two commonly used models in research and clinical settings, BCRAT and IBIS, in two large prospective cohorts.^8^ The current study aims to conduct an extensive independent validation of iCARE-based models that integrate classical risk factors and a 313-SNP PRS^17^ in 15 prospective cohorts across six countries. We also examined risk stratification of these models by estimating five-year risk projections in those countries. Finally, we provided projections for improved levels of risk stratification attainable by PRSs expected to result from ongoing efforts to double the size of current breast cancer GWAS.^20^

## METHODS

### Study Populations

Eligible person-time within the 15 cohorts began at the time of DNA collection for women of self-reported European ancestry aged 18-75 years with no personal history of breast cancer, who consented for genetic studies and completed a risk factor questionnaire. For studies with multiple questionnaires, we used those administered closest to DNA collection (Table S1). There was a wide range of variation in the year and average age of DNA collection across the populations (Table S1). The distribution of risk factors varied substantially across the cohorts (Tables S2-S3), across the underlying country-specific reference populations, and between cohorts and the reference population within the same country (Tables S4-S5).

### Risk Models

We used iCARE to build models for five-year absolute risk of developing breast cancer^19^ integrating classical risk factors and PRS (Figure 1). Age was used as the timescale in disease incidence modeling. The conditional age-specific incidence rates given the risk factors were assumed to follow a Cox proportional hazards model.^21^ Classical risk factors in the iCARE-Lit model were specified as previously described.^8^ Briefly, this model includes ages at menarche and first birth, parity, height, alcohol intake, breast cancer family history (i.e., presence or absence of disease in at least one first-degree relative), history of benign breast disease, oral contraceptive use and BMI for all women. For women 50 years or older it additionally includes age at menopause, hormone replacement therapy (HRT) use, and current HRT type. The relative risks for these factors were obtained from a literature review separately for women younger and older than 50 years (Supplements).^8^ The 313-SNP PRS, recently described in Mavaddat et al.,^17^ was integrated into the iCARE-Lit model assuming a multiplicative joint association with disease risk, except for family history where we accounted for attenuation of the relative risk due to its correlation with PRS.

**Figure 1.**
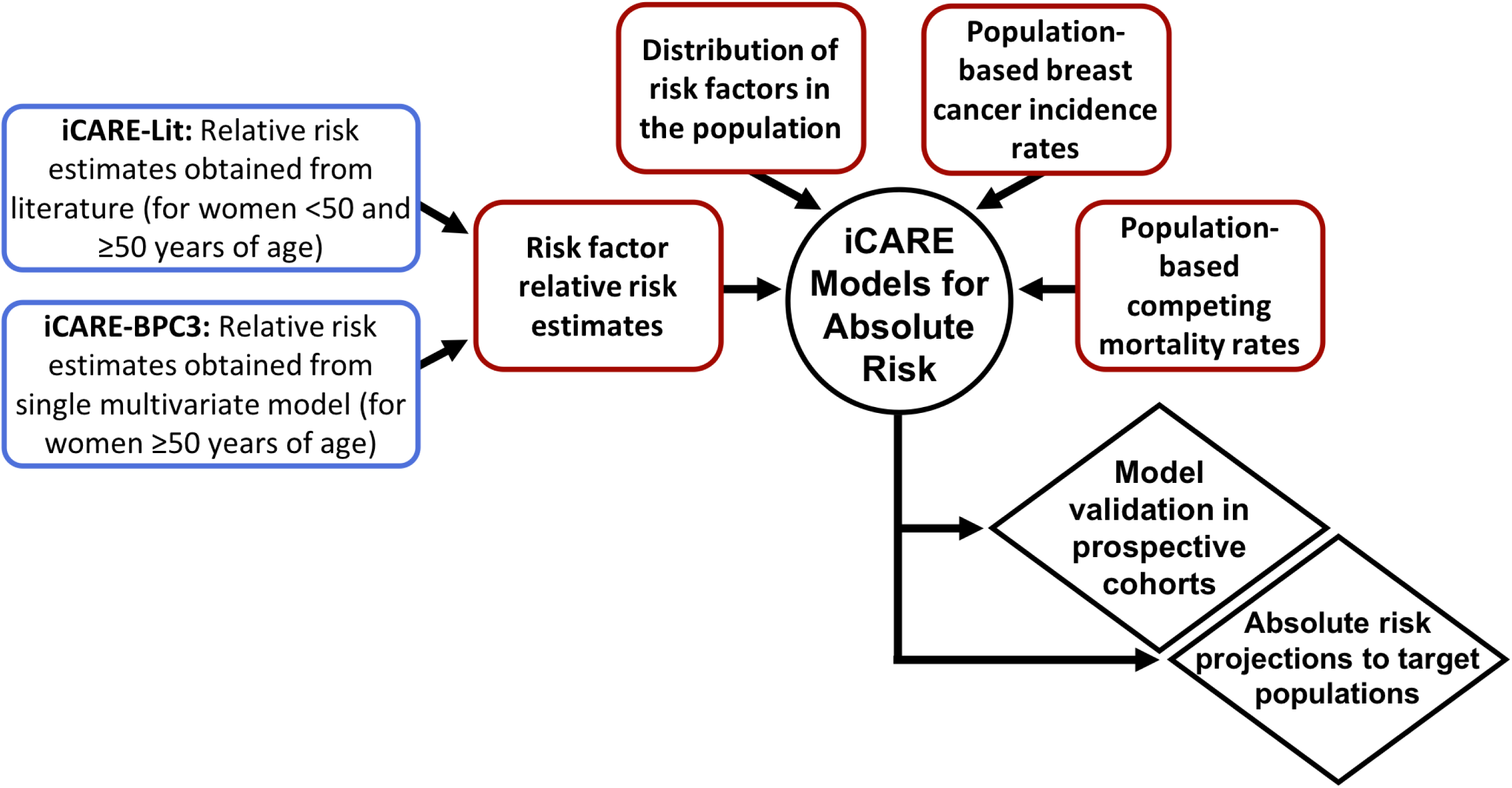
Conceptual diagram of model building and validation of Individualized Coherent Absolute Risk Estimator (iCARE) for breast cancer. iCARE-BPC3 = Individualized Coherent Absolute Risk Estimation model based on Breast and Prostate Cancer Cohort Consortium, iCARE-Lit = iCARE model based on literature review.

To estimate absolute risk of breast cancer, we incorporated country-specific incidence and competing mortality rates from population-based registries and the population distribution of risk factors, using an individual-level reference dataset representative of each population (Figure S1, Tables S4-S5). We simulated the PRS in the reference dataset assuming independence with classical risk factors, conditional on family history.^19^

### Cohort-Specific Validation of Risk Models

Women developing incident primary breast cancer, either in-situ or invasive, during the follow-up period were considered cases (Table S1). Follow-up for all subjects was assumed to begin one year following study entry to reduce the impact of prevalent cases on validation analyses and was defined up to the last contact or record linkage to cancer registry or five years, whichever came first. Predicted risk estimates account for competing mortality due to other causes during the follow-up period (Figure 1). The model validation methods are discussed in detail in our previous work^8^ and the Supplements.

### Meta-Analysis of Relative Risk Calibration

We evaluated relative risk calibration of each model through a meta-analysis across cohorts using the deciles of the relative risk score in UK Biobank as the standard set of cutoffs to delineate the risk categories. We first computed the relative risks (observed and expected) for each decile with respect to the fifth decile and combined the estimates using standard inverse variance weighted meta-analysis^22^ on the log-relative risk scale. We also computed a meta-analysis estimator and a Wald-statistic based 95% confidence interval (CI) of the AUC based on all risk factors except age.

### Projection of Risk to the General Populations

Projected five-year absolute risks for women of European ancestry aged 50-70 years in each country were calculated using the population distribution of ages and risk factors in the corresponding reference dataset, population-specific breast cancer incidence and competing mortality rates (Figure 1, Supplements). Proportions of women and incident cases expected to arise within five years were calculated at both extremes of the risk distribution based on two high-risk thresholds and two low-risk thresholds. Low-risk cutoffs were 0.6% and 1.13%, corresponding to average five-year risk for US women aged 40 and 50 years, respectively. High-risk cutoffs were 3%, corresponding to US Preventive Services Task Force recommendation for risk-lowering drugs^2^, and 6%, corresponding to a cutoff for very high risk used in the WISDOM trial.^23^ We also evaluated expected improvements in risk stratification by incorporating an improved PRS to the iCARE models (Supplements).

### Reclassification Analysis

We evaluated potential improvements in risk stratification achieved by adding genetic variants to the model with classical risk factors in the populations of the UK and the US in terms of proportions of women and future cases reclassified at the extremes of the risk distribution based on two low-risk (0.6% and 1.13%) and two high-risk cutoffs (3% and 6%). Women who were at 8% or higher risk, corresponding to the National Institute for Health and Care Excellence (NICE) guidelines for ten-year risk of a 40-year-old woman in the UK,^3^ were above 3% five-year risk of breast cancer.

## RESULTS

Meta-analyses of the iCARE-Lit model relative risk scores showed that observed and expected scores based on the 313-SNP PRS were not significantly different overall, with excellent calibration across risk deciles (Figure S2). While relative risk scores based on classical risk factors appeared well calibrated for most cohorts (Figures S3-S5), the meta-analysis showed 45% and 18% overestimation of risk for the highest decile for women younger and older than 50 years, respectively, and 16% underestimation of risk for women in the lowest decile, only in the older group (Figure S2). The integrated model also appeared well calibrated across cohorts (Figures S3-S5), and meta-analyses showed good calibration across most risk categories, but a 40% and 27% overestimation in the highest risk decile for the younger and older age groups, respectively (Figure 2, Figure S2). The analyses for women older than 50 years using a similar model with relative risk estimates from a multivariable analysis of prospective cohorts (iCARE-BPC3^24^) provided somewhat better calibration of models at the high-risk decile (Figure S2, Figure S6).

**Figure 2.**
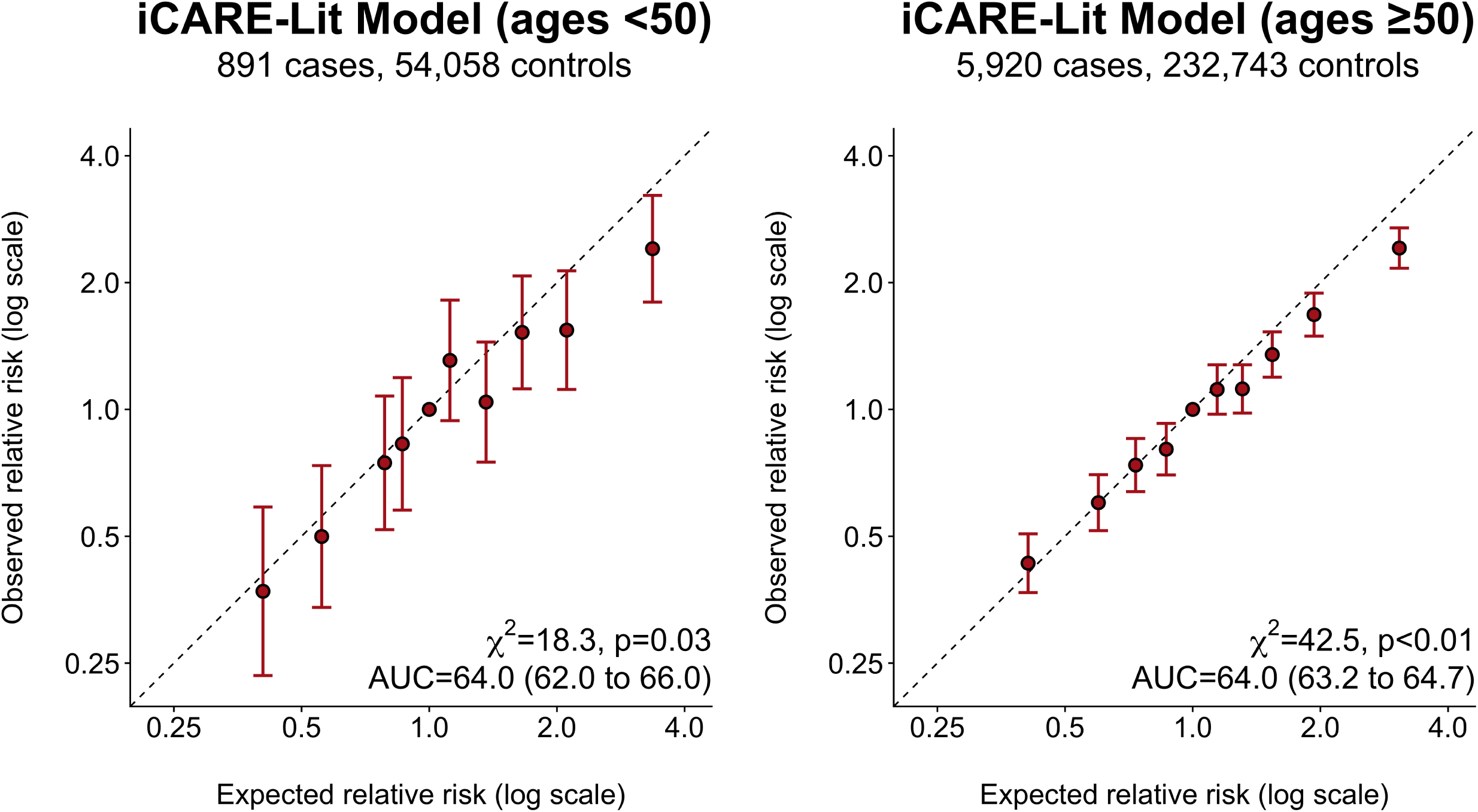
Relative risk calibration of integrated breast cancer risk models (with classical risk factors and PRS) based on meta-analysis across validation studies. Classical risk factors include age at menarche, age at first live birth, parity, oral contraceptive use, age at menopause, hormone replacement therapy use, type of hormone replacement therapy, alcohol intake, height, BMI, breast cancer family history (i.e., presence or absence of breast cancer in at least one first degree relative), and benign breast disease. Meta-analysis is based on a reduced set of risk factors that were available in the majority of the validation cohorts. History of benign breast disease and type of hormone replacement therapy (iCARE-Lit model for women 50 years or older) was set to missing for all subjects. Meta-analysis of the iCARE-Lit model for women younger than 50 years included GS, NHS II, and UK Biobank. Meta-analysis of the iCARE-Lit model for women 50 years or older additionally included CPS II, EPIC NL, EPIC UK, KARMA, MMHS, NHS, PLCO, and WGHS. The AUC estimates were adjusted for age at enrollment. Abbreviations: AUC = area under the curve, χ^2^ = chi-square test statistic, CPS = Cancer Prevention Study, EPIC = European Prospective Investigation into Cancer and Nutrition, GS = Generations Study, iCARE-Lit = iCARE model based on literature review, KARMA = KARolinska MAmmography Project, MMHS = Mayo Mammography Health Study, NHS = Nurses’ Health Study, PLCO = Prostate, Lung, Colorectal, Ovarian Cancer Screening Trial, PRS = polygenic risk score, UK = United Kingdom, WGHS = Women’s Genome Health Study.

For both age groups, incorporating the 313-SNP PRS to the classical risk factors substantially improved overall risk discrimination (Figure S2). Age-adjusted AUC (95% CI) was 55.9 (53.8 to 58.0) and 64.0 (62.0 to 66.0) for classical risk factors and integrated models, respectively, in younger women; and 57.3 (56.5 to 58.1) and 64.0 (63.2 to 64.7), respectively, for older women (Figure 2). However, the differences between the AUCs based on the 313-SNP PRS alone and integrated models were smaller, with overlap in 95% CIs (Figure S2). All the models showed improvements in AUC after incorporating the variation due to age at enrollment (Figure 3, Figures S11-S13).

**Figure 3.**
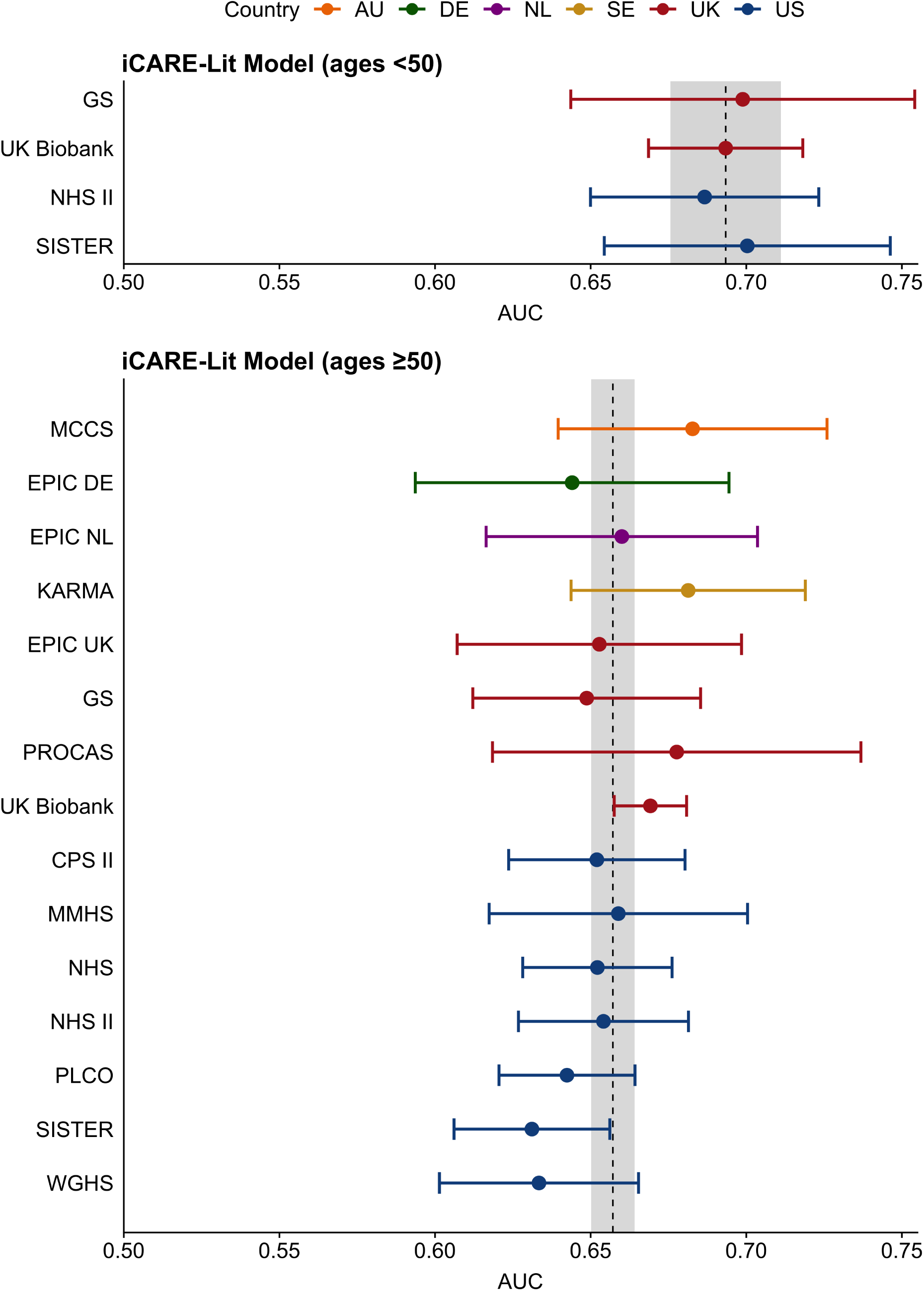
Cohort-specific AUCs for the integrated iCARE-Lit models (with age at enrollment, classical risk factors and PRS). Classical risk factors include age at menarche, age at first live birth, parity, oral contraceptive use, age at menopause, hormone replacement therapy use, type of hormone replacement therapy, alcohol intake, height, BMI, breast cancer family history (i.e., presence or absence of breast cancer in at least one first degree relative), and benign breast disease. The vertical dashed line and gray region represent the meta-analyzed AUC estimate and the 95% confidence interval for the meta-analyzed AUC. Abbreviations: AU = Australia, AUC = area under the curve, CPS = Cancer Prevention Study, DE = Germany, EPIC = European Prospective Investigation into Cancer and Nutrition, GS = Generations Study, KARMA = KARolinska MAmmography Project, MCCS = Melbourne Collaborative Cohort Study, MMHS = Mayo Mammography Health Study, NHS = Nurses’ Health Study, NL = the Netherlands, PRS = polygenic risk score, PLCO = Prostate, Lung, Colorectal, Ovarian Cancer Screening Trial, PROCAS = Predicting Risk Of Breast CAncer at Screening, SE = Sweden, UK = United Kingdom, US = United States, WGHS = Women’s Genome Health Study.

While calibration of absolute risk for the integrated models varied substantially across cohorts (Figures 4-5, S7A-S10B), the models did not systematically over- or under-predict risk. The median (range) expected-to-observed (E/O) ratio across the studies were 0.94 (0.72 to 1.01) for women younger than 50 years and 1.04 (0.73 to 1.31) for women older than 50 years. For women at the highest risk decile these were 0.9 (0.7 to 1.4) and 1.2 (0.7 to 1.6) for the two age groups, respectively.

**Figure 4.**
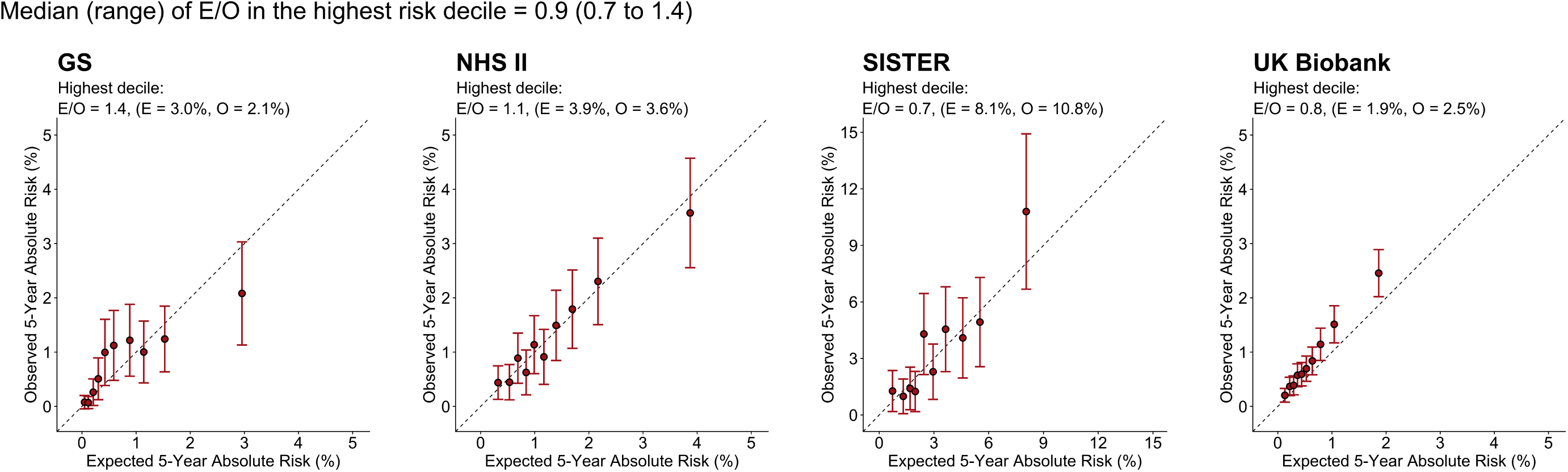
Absolute risk calibration for the integrated iCARE-Lit model with classical risk factors and PRS for women younger than 50 years (4 cohorts). Risk categories were defined based on deciles of predicted five-year absolute risk. Classical risk factors include age at menarche, age at first live birth, parity, oral contraceptive use, age at menopause, hormone replacement therapy use, type of hormone replacement therapy, alcohol intake, height, BMI, breast cancer family history (i.e., presence or absence of breast cancer in at least one first degree relative), and benign breast disease. Abbreviations: E = Average of predicted five-year risk in the highest decile of predicted five-year risk, GS = Generations Study, NHS = Nurses’ Health Study, O = observed proportion of subjects developing breast cancer in five years in the highest decile of predicted five-year risk, UK = United Kingdom.

**Figure 5.**
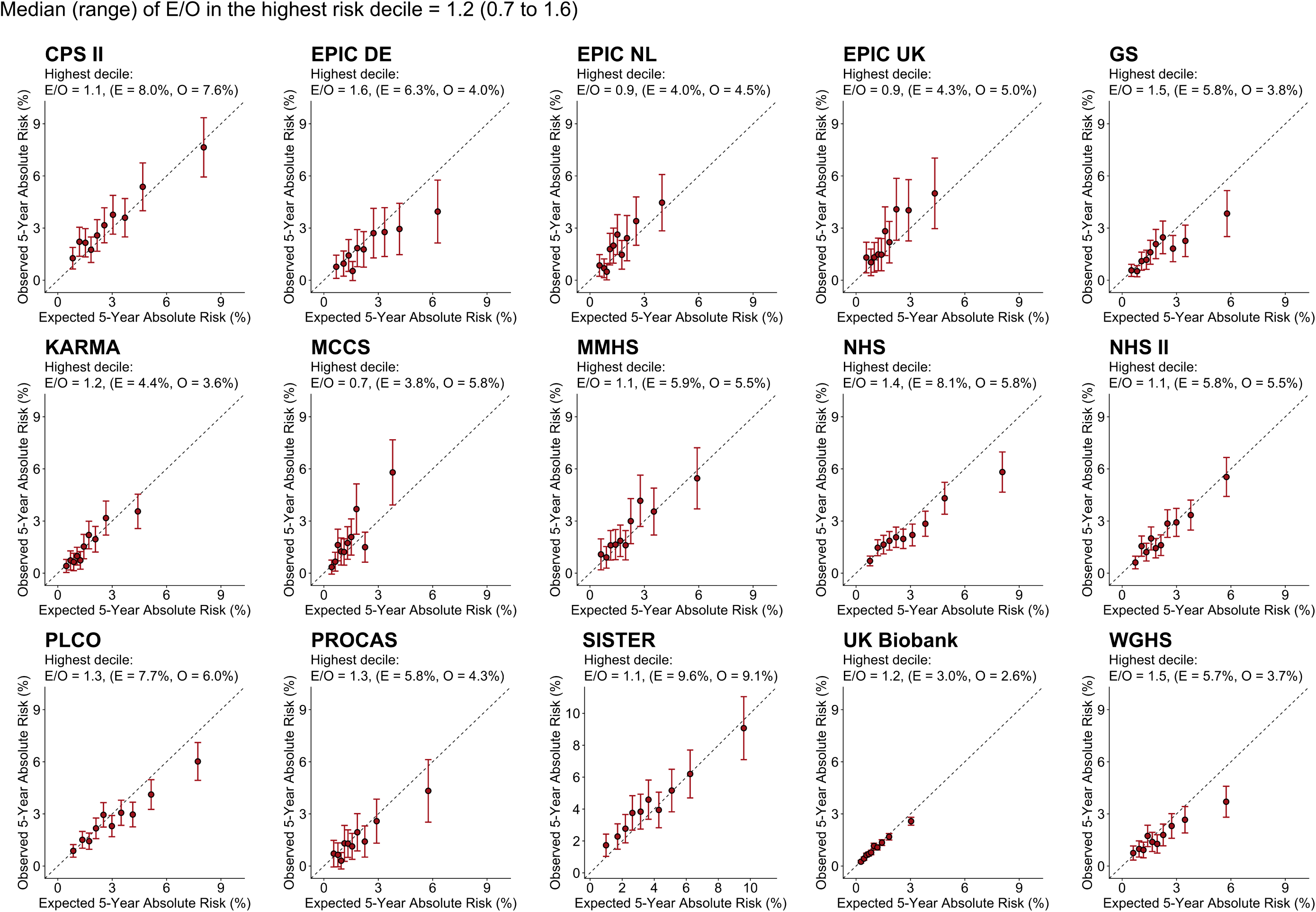
Absolute risk calibration for the integrated iCARE-Lit model with classical risk factors and PRS for women 50 years or older (15 cohorts). Risk categories were defined based on deciles of predicted five-year absolute risk. Classical risk factors include age at menarche, age at first live birth, parity, oral contraceptive use, age at menopause, hormone replacement therapy use, type of hormone replacement therapy, alcohol intake, height, BMI, breast cancer family history (i.e., presence or absence of breast cancer in at least one first degree relative), and benign breast disease. Abbreviations: CPS = Cancer Prevention Study, DE = Germany, E = Average of predicted five-year risk in the highest decile of predicted five-year risk, EPIC = European Prospective Investigation into Cancer and Nutrition, GS = Generations Study, KARMA = KARolinska MAmmography Project, MCCS = Melbourne Collaborative Cohort Study, MMHS = Mayo Mammography Health Study, NHS = Nurses’ Health Study, NL = the Netherlands, O = observed proportion of subjects developing breast cancer in five years in the highest decile of predicted five-year risk, PLCO = Prostate, Lung, Colorectal, Ovarian Cancer Screening Trial, PROCAS = Predicting Risk Of Breast CAncer at Screening, UK = United Kingdom, WGHS = Women’s Genome Health Study.

Risk projections among women of European ancestry aged 50-70 years showed that addition of PRS to the classical risk factors produced substantial improvements in risk stratification (Figure 6, Tables S7A-S7F), resulting in identifying a large fraction of the population at high or low risk (five-year risk >3% or ≤1.13%, respectively). These projections vary across populations due to differences in the distribution of age and classical risk factors. The proportion of women identified at low risk ranged from 27.0% in the US (∼8.1 of 30 million) to 46.0% in Germany (∼5.5 of 12 million). At the high-risk threshold, the proportion of women identified ranged from 7.0% in Germany (∼841,000 of 12 million) to 17.7% in the US (∼5.3 of 30 million). As a sensitivity analysis, we adjusted for the miscalibration of risk in the high-risk decile (Figure 2) by attenuating the risk projections by the amount of overestimation in that category. After this adjustment, the proportion of women identified at the high-risk threshold ranged from 5.6% in Germany (∼677,000 of 12 million) to 16.7% in the US (∼5 of 30 million) (Table S8). Proportions of future cases identified at high risk also remained substantial after adjusting for the relative risk miscalibration.

**Figure 6.**
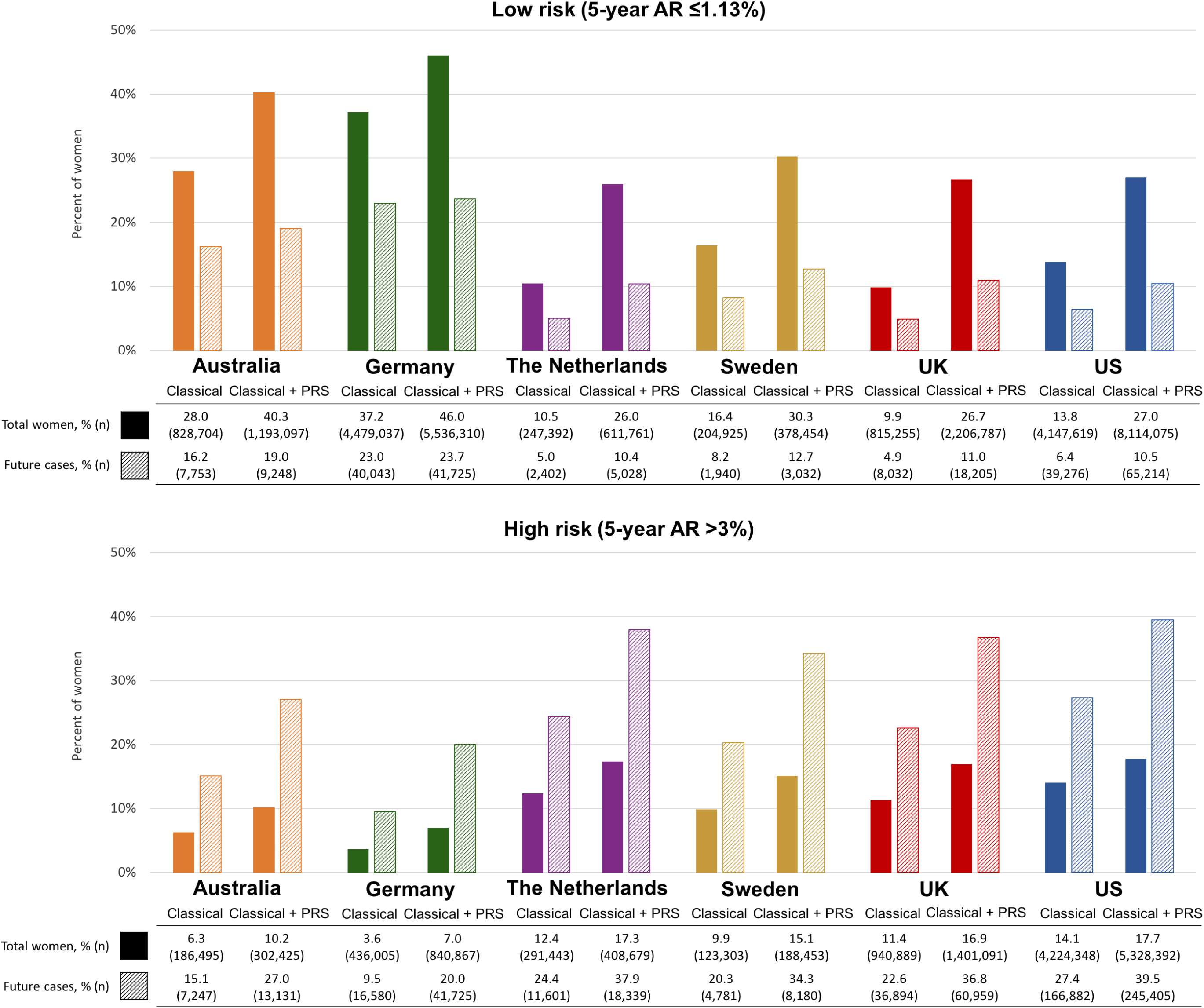
Women of European ancestry aged 50-70 years in the general populations of the six countries (Australia, Germany, The Netherlands, Sweden, the UK, the US) expected to be identified at low and high risk of breast cancer according to two risk thresholds and the incident cases of breast cancer expected to occur in these groups within a five-year interval. The expected number of women is calculated using 2017 population estimates (N = 2,960,506) from Australian Bureau of Statistics for Australia, 2016 population estimates (N=12,024,487) from the Federal Statistical Office for Germany, 2016 population estimates (N = 2,356,691) from the Central Agency for Statistics for the Netherlands, 2016 population estimates (N=1,249,695) from Statistics Sweden for Sweden, mid-2016 population estimates (N = 8,275,453) from the Office of National Statistics for the UK and mid-2016 population estimates (N = 30,030,821) from US Census Bureau for the US. The expected numbers of cases are estimated using the average predicted five-year risk in each population, calculated using the country-specific breast cancer incidence rates and risk factor distributions (Table S4). The 1.13% risk threshold corresponds to the average five-year risk of US women aged 50 years. The 3% threshold is used by US Preventive Services Task Force for recommending risk reducing medications. Classical risk factors correspond to the iCARE-Lit model and include age at menarche, age at first live birth, parity, oral contraceptive use, age at menopause, hormone replacement therapy use, type of hormone replacement therapy, alcohol intake, height, BMI, breast cancer family history (i.e., presence or absence of breast cancer in at least one first degree relative), and benign breast disease. Abbreviations: AR = Absolute risk, PRS = polygenic risk score, UK = United Kingdom, US = United States.

Our projections indicate that incorporating an improved PRS substantially increases the number of future cases expected to be identified in the high-risk group, with only a marginal predicted increase in total women in that group. In the low-risk group, an improved model would identify a substantially higher fraction of total women, with only a minimal number of additional cases expected (Tables S7A-S7F).

Addition of the 313-SNP PRS to the classical risk factors (Table 1, Table S6) resulted in a substantial reclassification of women leading to a net increase in the proportion of cases crossing risk thresholds under current clinical guidelines for breast cancer prevention. For example, for a five-year risk threshold of 3% recommended for risk-reducing medications in the US, 14.7% of US women were re-classified by the integrated model, leading to identification of an additional 12.3% of the future breast cancer cases. For the same threshold, the integrated model reclassified 15.3% of UK women, resulting in identifying an additional 15.7% of the future cases.

**Table 1.** Reclassification of women at high-risk thresholds after incorporating 313-SNP PRS to the classical risk factors. We report the number of women and future cases (i.e., women expected to develop breast cancer within five-years) above or below the risk threshold based on the classical risk factors and the number and percentage of these women moving below the threshold (down), above the threshold (up) and a total number and percentage of re-classified women. Classical risk factors correspond to the iCARE-Lit model and include age at menarche, age at first live birth, parity, oral contraceptive use, age at menopause, hormone replacement therapy use, type of hormone replacement therapy, alcohol intake, height, BMI, breast cancer family history (i.e., presence or absence of breast cancer in at least one first degree relative), and benign breast disease. The 3% cutoff corresponds to the US Preventive Services Task Force recommendation for risk-lowering drugs and the 6% cutoff corresponds to a cutoff for very high risk used in the WISDOM trial. Abbreviations: AR = absolute risk, PRS = polygenic risk score, SNP = single nucleotide polymorphism.

|  | UK population |  |  |  | US population |  |  |  |
| --- | --- | --- | --- | --- | --- | --- | --- | --- |
|  | 3% risk |  | 6% risk |  | 3% risk |  | 6% risk |  |
|  | Total women | Future cases | Total women | Future cases | Total women | Future cases | Total women | Future cases |
| **Based on classical risk factors only** |  |  |  |  |  |  |  |  |
| Above threshold, n (%) | 940,889<br>(11.4) | 34,955<br>(21.1) | 36,871<br>(0.4) | 2,407<br>(1.5) | 4,224,349<br>(14.1) | 169,055<br>(27.2) | 223,792<br>(0.7) | 16,327<br>(2.6) |
| Below threshold, n (%) | 7,334,564<br>(88.6) | 130,768<br>(78.9) | 8,238,582<br>(99.6) | 163,316<br>(98.5) | 25,806,471<br>(85.9) | 452,003<br>(72.8) | 29,807,028<br>(99.3) | 604,731<br>(97.4) |
| Total, N | 8,275,453 | 165,723 | 8,275,453 | 165,723 | 30,030,820 | 621,058 | 30,030,820 | 621,058 |
| **Based on classical risk factors + PRS** |  |  |  |  |  |  |  |  |
| Reclassified, n (%) | 1,265,899<br>(15.3) | 42,929<br>(25.9) | 140,656<br>(1.7) | 9,970<br>(6.0) | 4,424,697<br>(14.7) | 148,710<br>(23.9) | 790,733<br>(2.6) | 55,387<br>(8.9) |
| Moving down, n (%) | 402,848<br>(4.9) | 8,462<br>(5.1) | 17,753<br>(0.2) | 771<br>(0.5) | 1,660,327<br>(5.5) | 36,180<br>(5.8) | 108,699<br>(0.3) | 4,218<br>(0.7) |
| Moving up, n (%) | 863,051<br>(10.4) | 34,467<br>(20.8) | 122,903<br>(1.5) | 9,199<br>(5.5) | 2,764,370<br>(9.2) | 112,530<br>(18.1) | 682,034<br>(2.3) | 51,169<br>(8.2) |

## DISCUSSION

The integrated iCARE-Lit model with 313-SNP PRS and classical risk factors showed substantial improvements in breast cancer risk prediction across 15 prospective cohorts of women of European ancestry in six countries. The integrated models generally showed good calibration and provided wider stratification of population risk, identifying more women crossing clinically actionable risk thresholds used by current guidelines for breast cancer prevention and early detection.

The integrated iCARE-Lit model provided well-calibrated relative risk scores across validation cohorts (Figures S3-S5); however, meta-analyses showed some over-prediction in the highest risk decile that was driven by the classical risk factor component. The calibration of absolute risks, however, varied more widely across cohorts, even within countries, though we did not see evidence of systematic under- or over-prediction across studies. This suggests that differences across cohorts are likely due to random variation or differences between study populations (e.g., wide range of study time periods (1989-2013) and differences in risk factor distributions or disease rates), rather than a reflection of intrinsic model properties. This highlights the importance of absolute risk validation across multiple study populations, particularly using cohorts similar to the target populations, both in chronologic years of study and underlying risk. Further studies in countries represented here by only one cohort, or not included in this report, are needed to evaluate country-specific differences in model performance.

We recently showed that five-year risk predictions by the iCARE-Lit model based on classical risk factors were at least as accurate as two established models used in clinical practice: BCRAT (“Gail”) and IBIS (“Tyrer-Cuzick”).^8^ Previous evaluations (e.g., Terry et al. 2019^6^) of BCRAT and IBIS also showed overestimation of risk in the highest risk categories.^25-27^ Our analyses suggested that building models from multivariable analysis of classical risk factors in prospective cohorts^24^, rather than from the literature, could improve calibration at the high-risk decile. Thus, calibration and discrimination of models can potentially improve through efforts of building multivariate relative risk models in prospective cohorts of women across age groups, and with more comprehensive information on questionnaire-based risk factors. Addition of risk biomarkers such as mammographic breast density,^11,12,27,28^ circulating hormone levels,^28-30^ or novel risk factors as they are identified in the future can result in further improvements.

Discriminatory accuracy of risk models may be substantially different in research cohorts than in target populations due to differences in underlying risk factor distributions. For instance, our projections show a higher discriminatory accuracy in the US population (AUC = 66.5, Table S7F, Figure S14) compared to the US-based cohorts (AUC range: 63.1-65.8) (Figures S11-S12). Our projections also show that an improved PRS, achievable through larger GWAS, could lead to better risk stratification, with a model integrating risk factors and an improved PRS achieving an AUC∼0.71 (Tables S7A-S7F). This will improve our ability to identify women eligible for risk-reducing interventions, or supplemental screening by magnetic resonance imaging or other imaging modalities. However, since the discriminatory performance of models will remain moderate, most breast cancers will still occur among women not identified at elevated risk. Thus, broader public health efforts targeting the whole population will continue to be required for reducing the population burden of breast cancer in a major way.^31^ Risk-stratified screening strategies at the population level tailored to women’s individual risks based on integrated models may improve the effectiveness of population-based screening, relative to the current age-stratified programs,^32^ and is currently being evaluated in screening trials^33^.

We used country-specific breast cancer incidence rates and a reference dataset for each country, built from population-based surveys, to translate relative risks to absolute risk estimates over a specified time period.^19^ This critical step in building absolute risk models minimizes miscalibration of absolute risk when models developed for one target population are applied to other populations or countries. Although most model algorithms allow for changes in default incidence rates, the underlying risk factor distributions are often implicit and cannot be easily changed. While this flexibility is a strength of the iCARE modeling approach, it requires the availability of population-based survey data with information on all risk factors included in the models for the relevant time periods. We were unable to identify a single data source for the distribution of all the risk factors in each target population, requiring us to simulate some risk factors and make various modeling assumptions (e.g., independence of certain risk factors). This could have affected model performance across study populations. Finally, the reference datasets enable iCARE models to provide absolute risk estimates for individuals based on a subset of risk factors in the model.^19^ This feature adds flexibility to use the models in different settings using information on a subset of risk factors.

Our risk models are aimed at the general population and do not adequately capture risk for women with strong family histories or carrying high-risk mutations. This requires integration with family-based models, e.g., our recent extension of the BOADICEA model^13^ to include the iCARE-Lit risk factor component. However, this fully extended model has not yet been prospectively validated. Although iCARE can be used for risk predictions over any time period, the current study only evaluated five-year risk prediction, and further work is needed to evaluate longer-term predictions used by some clinical guidelines.

The iCARE-Lit model includes parameters for atypical hyperplasia, lobular carcinoma in situ and other benign breast diseases. However, this information was not available from the participating cohorts, thus further validation is required for risk prediction for women with these conditions. The iCARE models predict risk of overall breast cancer (i.e., invasive and *in situ*), rather than specific subtypes. Because risk factor associations and the effectiveness of preventive and screening strategies vary by tumor subtypes (e.g., estrogen receptor positive and negative tumors),^34-39^ future work on subtype-specific risk predictions could result in more precise identification of women who would benefit most from specific interventions. Finally, the current models were derived and evaluated in studies of women of European ancestry and additional studies are urgently needed to develop and validate models for other populations, for whom alternative models have only been evaluated in relatively small studies.^40-42^

In summary, we present extensive validation results of a breast cancer risk prediction model integrating a newly developed PRS and classical risk factors. We show that it can provide substantial improvement in risk assessment for application of current clinical guidelines, or future risk-stratified prevention and screening strategies.^43,44^

## Data Availability

N/A

## AUTHOR CONTRIBUTIONS

MGC and NC conceived the study. PPC developed and implemented the method and code for the statistical analyses. ANW coordinated analyses with the lead analysts from each cohort to obtain results. CG, AH, ME, MS, CS, BDC, KM, and EH analyzed data to generate preliminary results from the participating cohorts. ANW analyzed preliminary results from each study to generate the final version of the results presented in the main manuscript and supplementary materials. ANW, PPC, MGC and NC interpreted the findings and led the writing of the manuscript. All authors contributed to the writing and reviewing of the final manuscript and its earlier draft.

